# Global Lipidomic and Metabolomic Uncovers Blood Signatures of Left Ventricular Assist Devices for Heart Failure

**DOI:** 10.1101/2024.11.19.24317588

**Authors:** Na Zhang, Hao Chen, Heping Li, XiaoYu Xu, Xuman Zhang, Haitao Hou, Zhifu Han, Guowei He, Yu Zhang

**Affiliations:** Functional test platform of implanted and interventional cardiovascular devices,School of Basic Medical Sciences,Tsinghua University, Beijing 100084,China; School of Clinical Medicine,Tsinghua University, Beijing 100084,China; Department of Engineering Physics, Tsinghua University, Beijing 100084, China; Academy of Arts & Design, Tsinghua University, Beijing 100084, China; ROCOR Medical Technology Co., Ltd, Tianjin TEDA 300457,China; TEDA International Cardiovascular Hospital, Tianjin University, 61.Third Avenue. TEDA 300457, China; Tianjin Key Laboratory of Molecular Regulation of Cardiovascular Diseases and Translational Medicine, Tianjin, China

**Keywords:** LVAD, heart failure, lipidomic, metabolomic;molecular mechanisms

## Abstract

**Background:** The left ventricular assist device (LVAD) significantly improves the health of patients with chronic advanced heart failure (HF); however, its underlying molecular mechanisms remain unclear. This study aimed to develop an integrated plasma pseudo-targeted lipidomic and untargeted metabolomic strategy to provide insight into the early postoperative changes that occur in the global blood metabolome profile and determine whether these changes can be used to screen patients for LVAD installation.

**Methods:** Data was collected from 20 pairs of patients with HF before and after LVAD surgery and compared with 36 healthy subjects. Plasma metabolomic and lipidomic profiles were established by liquid chromatography-mass spectrometry and analyzed by multivariate statistics.

**Results:** A total of 49 lipids showed significant recovery after LVAD pump loading compared with before pump loading. Moreover, 144 differential metabolites and 21 pathways were identified from healthy control and patients with HF. Among which, 33 metabolites were differentially regulated between pre and post-LVAD samples (p < 0.05, FC > 2). Further analysis revealed differential regulation in two key pathways: fatty acid metabolism and methionine metabolism. Simultaneously, we identified S-adenosylmethionine, L-methionine, FFA (14:1), and FFA (16:1) as potential diagnostic markers for the prediction of LVAD efficacy in HF. In three postLVAD patients who died within one year, we observed a decrease in SM (24:0) and SM (22:0) immediately before LVAD implantation, indicating that these metabolites may predict a poor outcome. Furthermore, we demonstrated that PS (18:1/20:4) and canavaninosuccinate were significantly attenuated in postLVAD patients.

**Conclusions:** Our findings provide preliminary evidence that LVAD therapy is associated with changes in the metabolomic and lipidomic profiles of patients with HF. It highlights the potential use of metabolomics as a tool to stratify LVAD patients based on the risk of adverse events. These findings may help to guide patient selection for advanced HF therapies and identify new HF therapeutic targets.

**Graphical Abstract:** 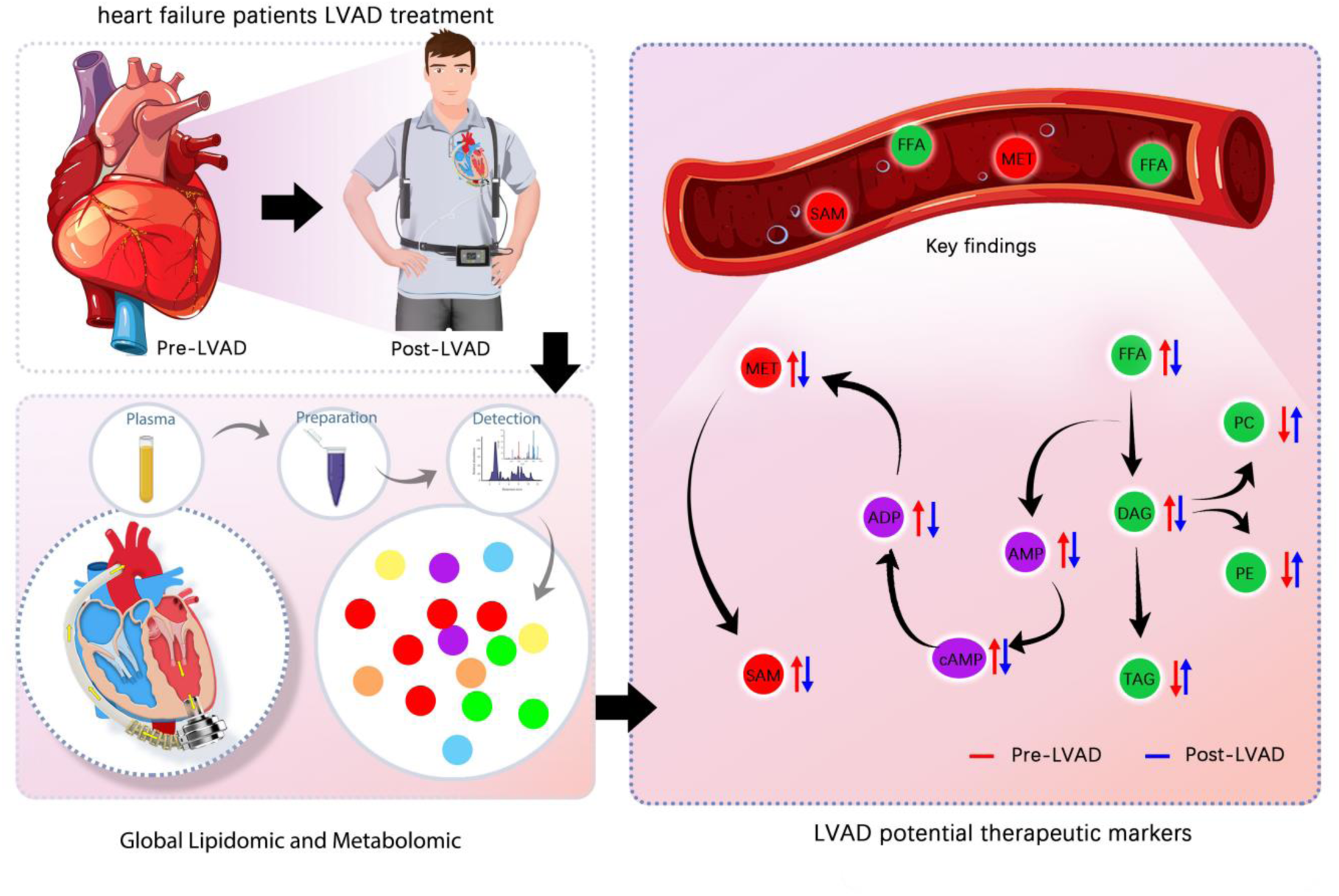

## Introduction

Heart failure (HF) is a significant contributor to morbidity and mortality worldwide with a five-year mortality rate of approximately 50% ^[1, 2]^. More than 64 million people with HF worldwide, form the so-called “HF pandemic.” ^[3]^ Left ventricular-assist devices (LVADs) are currently used as a nonpharmacologic treatment for patients with advanced HF ^[4]^. The implantation of LVADs has traditionally served as either a temporary bridge until cardiac transplantation or, because of the limited availability of donor hearts, as a lifetime destination therapy ^[5–7]^. Patients with LVAD support have an increased first-year survival rate and an overall improved quality of life ^[8–11]^. Compared with drug therapy, mechanical support improved symptoms more quickly and effectively in patients with advanced disease. Studies have demonstrated that LVAD implantation recovers myocardial function and regulates adverse metabolic remodeling in a subset of patients with HF ^[12–15]^. While the association between LVAD placement and the improvement of symptoms in patients with HF is well-documented, the mechanism underlying the effects of LVAD is poorly understood.

A metabolomics approach to LVADs is promising as it has been applied extensively to study the mechanisms of disease and drug action ^[16–18]^. Lipidomics is the systematic analysis of all lipids and relevant lipid pathways ^[19]^ and is a subfield of metabolomics. Lipids are recognized not only as a source of energy and as essential cellular components involved in organelle homeostasis and cell signaling, but also as regulators of interorgan communication and metabolism ^[20]^. Numerous studies have indicated that the metabolic dysregulation of lipids contributes to the etiology of HF. Therefore, screening for potential lipid biomarkers may contribute to the comprehensive evaluation of the mechanism of LVADs for HF treatment.

In our previous research, we initially reported a new LVAD called Heartcon, the implantation of Heartcon was a safe and useful LVAD for treatment of end-stage HF ^[21]^. In this study, we developed an integrated plasma pseudo-targeted lipidomic and untargeted metabolomic strategy to analyze the global blood metabolism characteristics of Patients with HF undergoing our LVADs therapy. Our primary goal was to determine which metabolites changed as a consequence of LVAD placement and the secondary aim was to determine whether these changes could help screen patients before LVAD installation. To our knowledge, this is the first report of the complete metabolomic and lipidomic analysis of LVAD-assisted HF patient plasma. This integrated strategy may provide a method to analyze the mechanism of action of medical equipment in the human body.

## Methods

### Participants and study design

A total of 76 plasma samples were collected from 56 subjects and divided into three groups: Patients with HF (preLVAD, n = 20; postLVAD, n = 20, paired samples) and healthy controls (HCs, n = 36). Patients with dilated ischemic heart failure were recruited from the Aerospace Taixin Group (Beijing, China) and diagnosed based on the Heart Failure Society of America criteria. HCs were recruited from volunteers for routine health examinations at Tsinghua University Hospital (Beijing, China).

### Biospecimen collection and processing

Venous blood from fasting subjects was collected into EDTA vacutainers by venipuncture. Plasma was isolated, transferred to 1.5 mL microtubes, centrifuged at 3,000 rpm for 5 min, and stored at -80°C. The experimental samples were prepared as follows: 100 μL of plasma was deproteinized with four volumes of methanol: chloroform (2:1) containing internal standards (ISs, Table S1), vortexed for 3 min, and kept at 4°C for 30 min. After centrifugation at 13,000 rpm for 15 min, the upper lipid layer and the lower polar layer solutions were transferred separately to a centrifuge tube. The upper solution was centrifuged at 13,000 rpm for 15 min for metabolomics analysis. The lower solution was concentrated and dried. The dried samples were reconstituted in 50 μL of methanol for lipidomics analysis. To evaluate the reproducibility and quality of the procedure, a quality check was done through the identical preparation and running of quality control (QC) samples made by mixing equal amounts of each sample.

### Lipidomics analysis

Lipidomics analysis was done using an ACQUITY Ultra Performance Liquid Chromatography instrument coupled with a SCIEX 6500 Triple Quadrupole mass spectrometer (UPLC-QQQ-MS, SCIEX, Milford, MA, USA). Extracts were retained and gradient eluted from an ACQUITY UPLC BEH C8 column using 0.1% acetonitrile in water and 0.1% acetonitrile formate in ESI^+^ and ESI^-^ mode. Detailed chromatographic and MS conditions are provided in the supplementary materials.

### Metabolomics analysis

Metabolomics analysis was performed using ACQUITY Ultra Performance Liquid Chromatography coupled with a Snapt G2-Si QTOF mass spectrometer. (UPLC-QTOF-MS, Waters, Milford, MA, USA). Extracts were retained and gradient eluted from an ACQUITY UPLC BEH HILIC column in 10% water and 90% acetonitrile containing 10 mM ammonium formate as solvent A and 50% water and 50% acetonitrile containing 10 mM ammonium formate as solvent B in ESI^+^ and ESI^-^ mode. Detailed chromatographic and MS conditions are provided in the supplementary materials. Instrument control and data acquisition were conducted using MassLynx 4.2 software.

### Raw data preprocessing

Total ion chromatograms for the metabolome data were imported into Progessis QI software for peak extraction and alignment (version 2.0, Waters). Total ion chromatograms for the lipid data were analyzed using Progessis QI software (version 2.0, Waters). Metabolomic features observed in at least 80% of the samples within the HF or control groups were retained. Original datasets were calibrated with ISs before statistical analysis. Similarly, ion features in the QC samples were also calibrated with ISs. The relative standard deviation was calculated after calibration.

### Metabolite identification

Metabolite identification was done using QI software, the HMDB database, together with an in-house library that included 1,000 metabolites involved in 15 pathways using UNIFI software. Comprehensive procedures scores were applied to remove the artifacts and background noise, including mass errors, fragments, and database searches.

### Statistical analysis

Multivariate analyses, including principal component (PCA), partial least square discriminants (PLS-DA), and orthogonal PLS-DA (OPLS-DA), were performed using SIMCA-P software (version 14.0, Umetrics, Umea, Sweden) and the MetaboAnalyst 3.0 online resource (https://www.metaboanalyst.ca/). A permutation test was conducted to check for overfitting. Volcano plots were applied to identify differential metabolites. Heat map analysis was performed using MetaboAnalyst. Correlation analysis was done using Origins 4.0 software (OriginLab, America). The predictive performance of the model was evaluated by the receiver operating characteristic curve (ROC) and area under the curve (AUC) parameters, which are generally used to determine overall discriminant ability.

## Results

### Study participants and experimental design

An overview of the main comparison groups is shown in **Figure 1** with details listed in **Table 1**. A total of 56 individuals were enrolled and 76 samples were divided into three groups. The control group included 21 females and 15 males with an average age of 36 ± 14 years. None of the control patients had been diagnosed with cancer, chronic inflammatory disease, chronic obstructive pulmonary disease, or were immunosuppressed. The HF group included 20 patients (average age 50 ± 13 years, 5 women and 15 men) with dilated cardiomyopathy on NYHA functional class IV and INTERMACS class 2. Two types of samples were collected from the HF group, before LVAD implantation (preLVAD) and 30 days after LVAD implantation (postLVAD). All patients stayed in the hospital for one month following LVAD implantation. Their diet and medication were regulated to avoid any effect on the experiment. In addition, functional and morphological parameters, such as left ventricular ejection fraction (LVEF) and B-type natriuretic peptide (BNP), were measured before and after LVAD implantation.

**Figure. 1.**
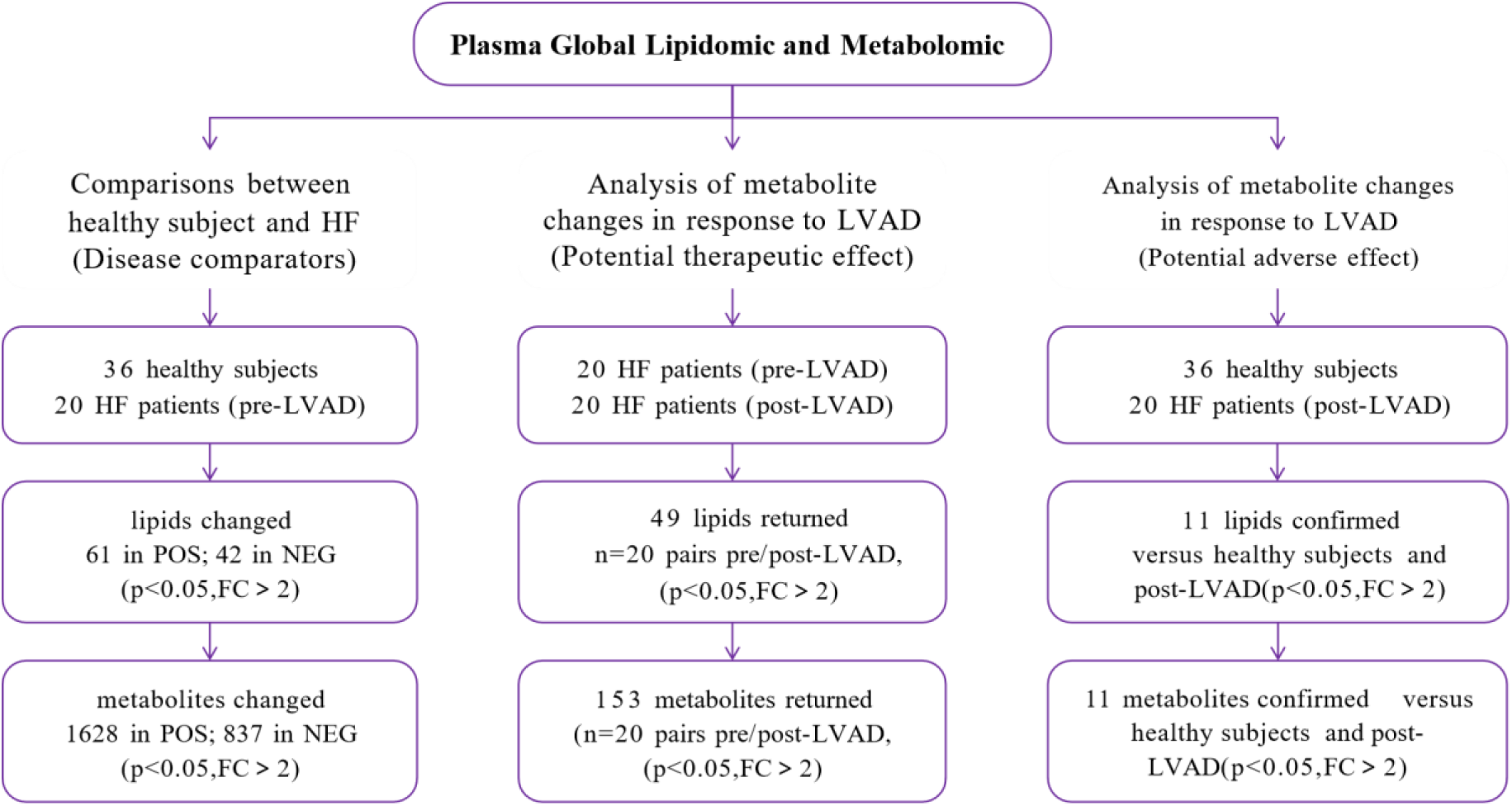
Main analyses study flowchart and characteristic metabolites identified in the main analyses

**Table 1.**
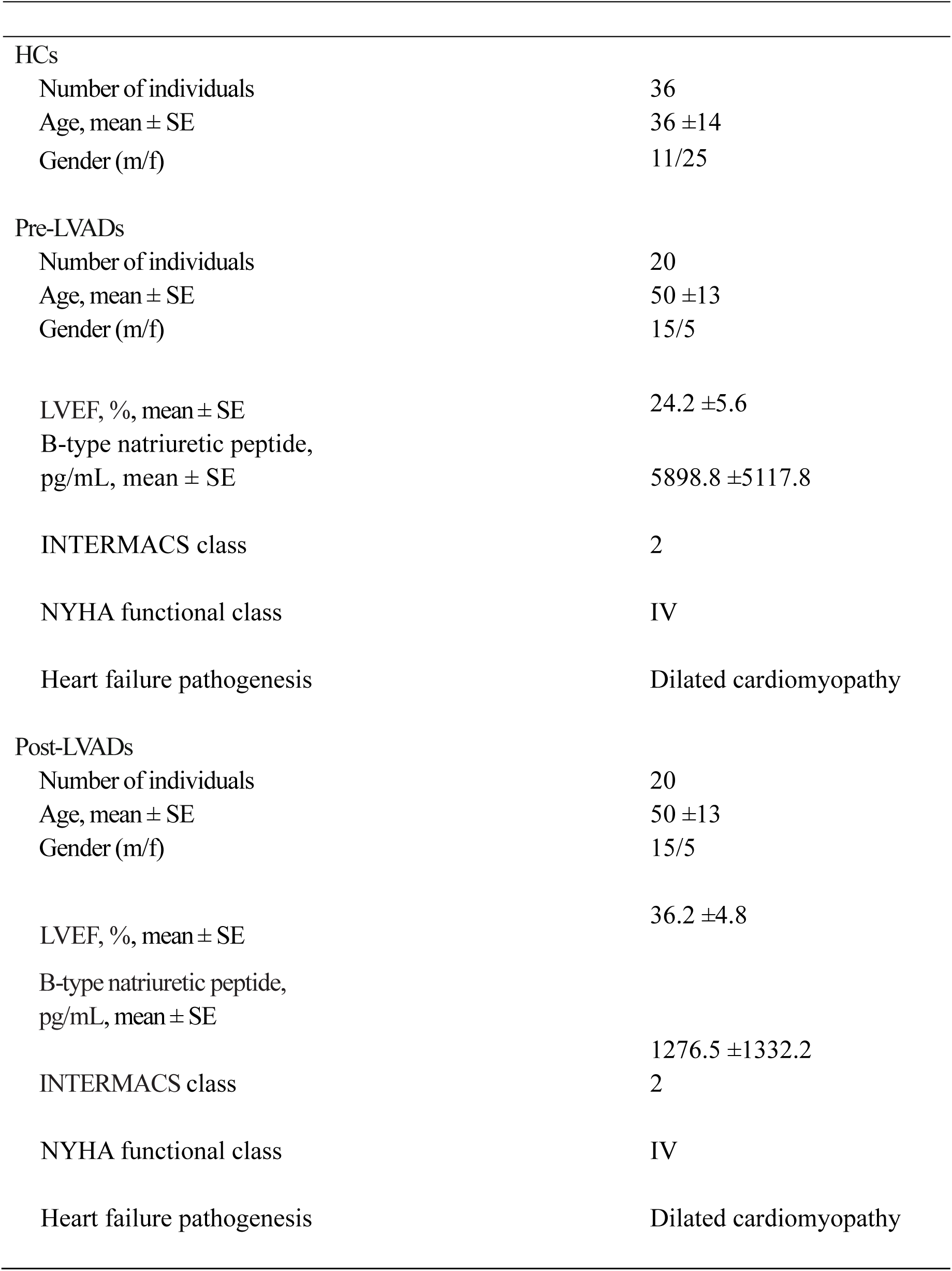
Study population features.

|  |  |  |
| --- | --- | --- |
| HCs |  |  |
| Number of individuals |  | 36 |
| Age, mean ± SE | | 36 ± 14 |
| Gender (m/f) |  | 11/25 |
| Pre-LVADs |  |  |
| Number of individuals |  | 20 |
| Age, mean ± SE | | 50 ± 13 |
| Gender (m/f) |  | 15/5 |
| LVEF, %, mean ± SE | | 24.2 ± 5.6 |
| B-type natriuretic peptide,<br>pg/mL, mean ± SE | | 5898.8 ± 5117.8 |
| INTERMACS class |  | 2 |
| NYHA functional class |  | IV |
| Heart failure pathogenesis |  | Dilated cardiomyopathy |
| Post-LVADs |  |  |
| Number of individuals |  | 20 |
| Age, mean ± SE | | 50 ± 13 |
| Gender (m/f) |  | 15/5 |
| LVEF, %, mean ± SE | | 36.2 ± 4.8 |
| B-type natriuretic peptide,<br>pg/mL, mean ± SE | | 1276.5 ± 1332.2 |
| INTERMACS class |  | 2 |
| NYHA functional class |  | IV |
| Heart failure pathogenesis |  | Dilated cardiomyopathy |

### LVAD improves fatty acids metabolism disorder in Patients with HF

We determined the differences in lipid distribution in patients with HF, before and after loading the pump, using a pseudo-targeted lipidomic method. The lipid metabolism characteristics of the HF group postLVAD pump were compared with those of normal individuals. A total of 736 lipids (365 and 371 lipids in positive and negative ion mode, respectively, Table S1 and Figure S1) with the highest intensities at their optimal collision voltages were selected to establish a final MRM transition list for the pseudo-targeted analysis. We first assessed the quality of the features drawn from the lipidomic analysis. The QC samples clustered in the center on the principal component analysis (PCA) score plots, suggesting that the analyses were reproducible and robust (Figure S2).

Using pseudo-targeted lipidomic data, we determined how LVAD placement improved the lipid metabolism of Patients with HF (**Figure 2A–E**). There were lipidome dissimilarities among different groups (as measured by partial least squares discriminant analysis, PLS-DA, **Figure 2A, B**). Orthogonal partial least squares discriminant analysis (OPLS-DA) was performed between Patients with HF before and after pump installation and between healthy subjects and the preLVAD HF group (Figure S3A–D). A permutation test was done to ensure that the model was not overfitted (Figure S3E–F). A volcano plot analysis revealed significant differences in 124 lipids between the HF and HC groups, whereas 164 lipids differed significantly between the preLVAD and postLVAD Patients with HF, with an FDR > 2 and *p* < 0.05 (Figure S4A–F).

**Figure. 2.**
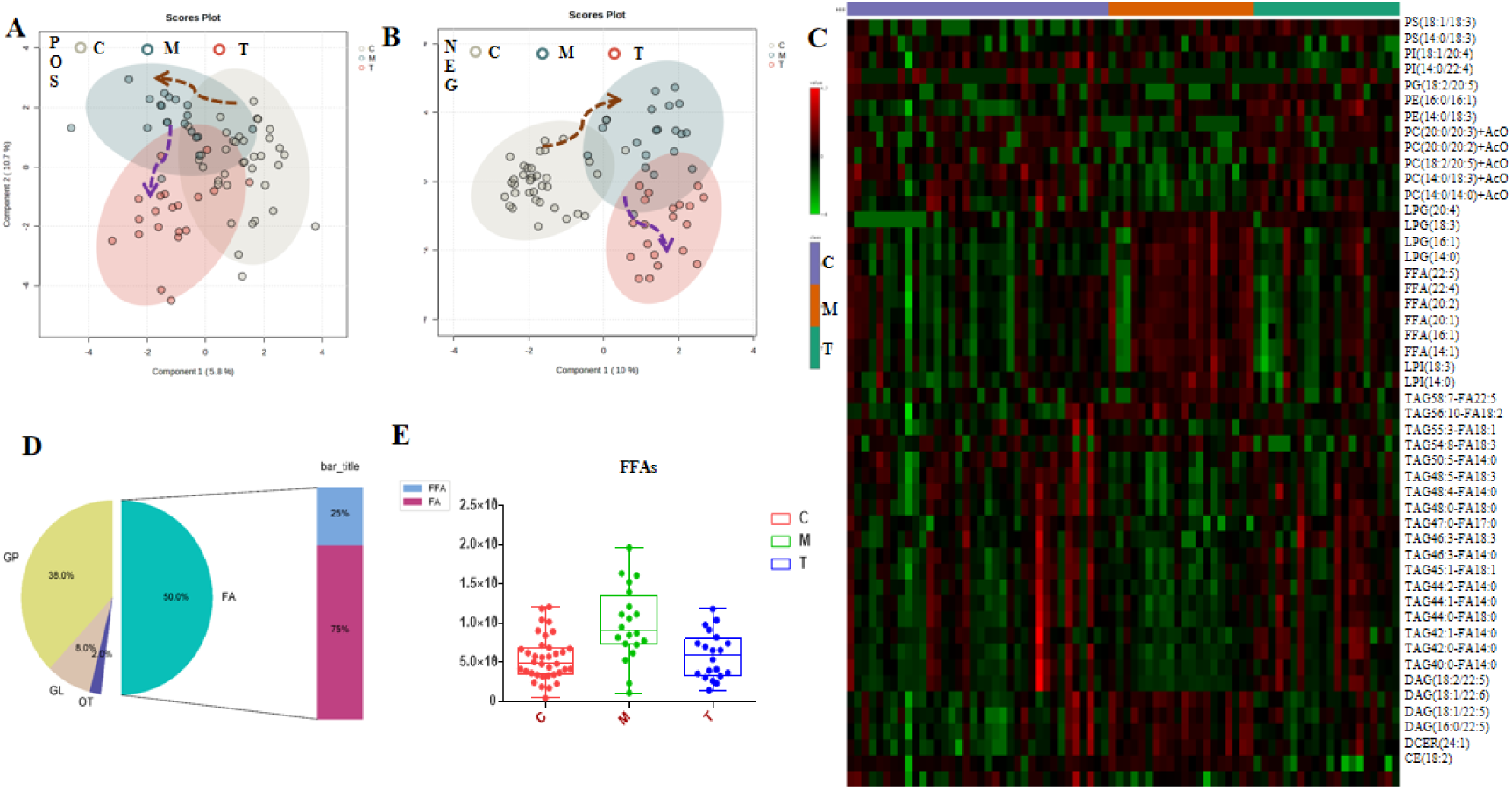
The positive effect of LVAD placement on the plasma lipid metabolism (a-b: PLS-DA analysis in positive and negative mode; c: lipid heat maps associated with the potential effect markers of LVAD; d: classification of potentially active lipid markers; e: box diagram of typical markers FFAs)

To determine the potential therapeutic mechanisms of LVAD treatment in HF, we assessed the intersecting differential lipid profiles using a diagram analysis (Figure S4C, F). A total of 49 lipids showed significant recovery postLVAD compared with that in preLVAD patients (**Figure 2C**). These 49 lipids included PS (18:1/18:3), PI (18:1/20:4), PG (18:2/20:5), PE (16:0/16:1), PC (20:0/20:2), LPG (16:1), FFA (20:1), LPI(18:3), TAG 44:0-FA18:0, DAG (18:1/22:6), and CE (18:2). These results indicate that fatty acids (FAs) are primarily associated with the therapeutic mechanism of LVAD (**Figure 2D**). Interestingly, free FAs (FFAs) were markedly increased in Patients with HF, which significantly decreased following pump installation. In contrast, FAs markedly decreased in the preLVAD group, whereas they were significantly increased in the postLVAD group (**Figure 2C**). To quantitate this shift in FAs and FFAs, we used box plots based on mass spectrometry abundance to visualize the differences (Figure S5). The box plot of the most remarkable substances, FFAs, is shown in **Figure 2E**. The results indicated that the abundance of FFAs was markedly increased after HF, but immediately restored to control levels with LVAD after one month.

Changes in FFAs through the LVAD procedure were also highlighted in a paired Wilcoxon analysis of the individual participants (Table S2). The efficiency of the six FFAs identified in 20 Patients with HF was 81%. Five lipids, including FFA (14:1), FFA (16:1), FFA (20:2), FFA (22:4), and FFA (22:5), were significantly improved between preand postLVAD, with a recovery rate of approximately 80%. Only FFA (20:1) was less than 80% efficient as it failed to improve following the LVAD procedure in five patients. FFAs showing marked improvement after LVAD implantation may be considered potential efficacy markers. As an example, we showed paired bar charts for FFA (22:4) (**Figure 3**).

**Figure. 3.**
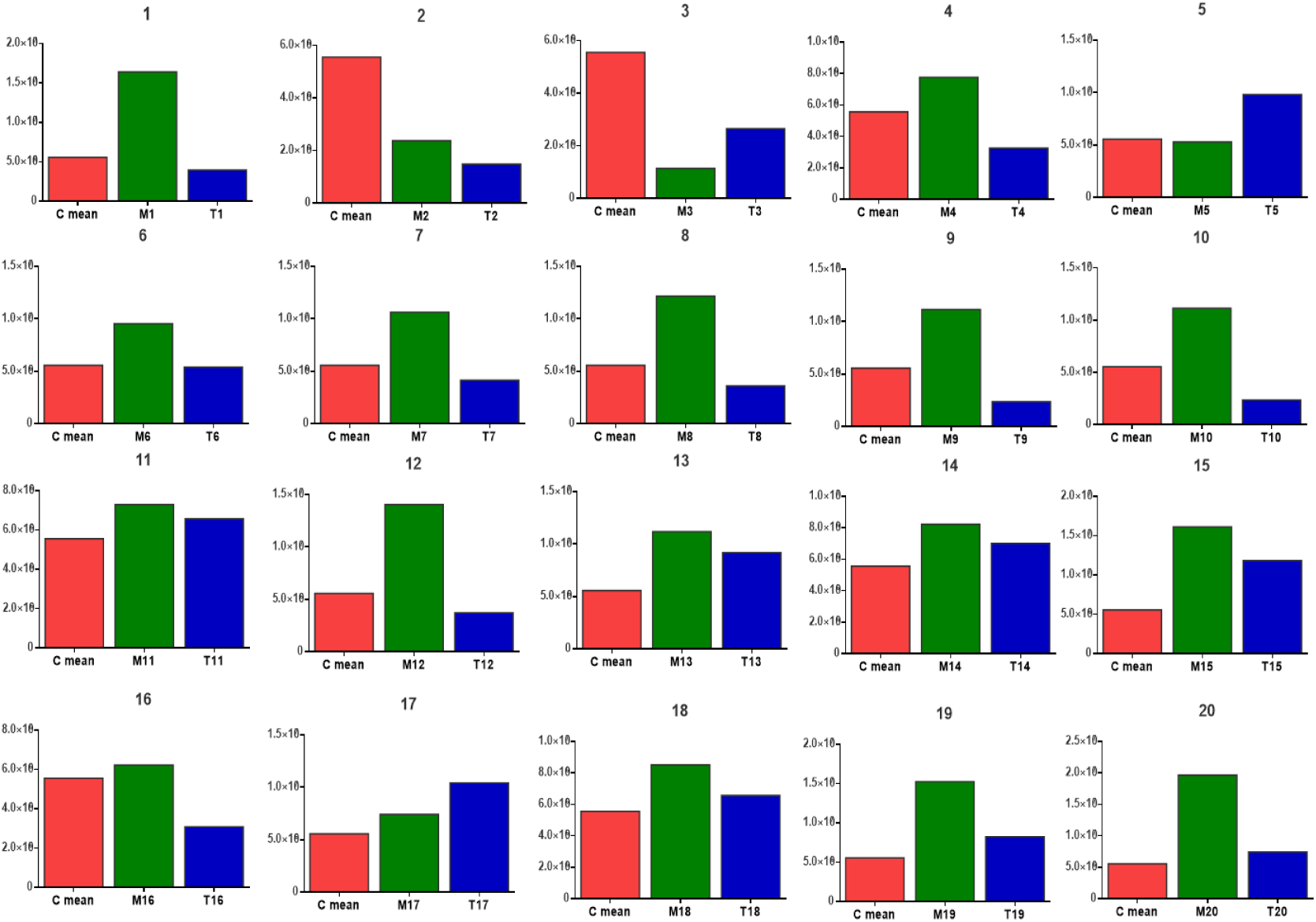
Semi-quantitative analysis of FFA (22:4) as an example of FFAs with potential therapeutic markers in 20 HF individuals

### LVAD improves methionine metabolism disorder

There was a surprising improvement in the homeostasis of small polar lipid metabolites postLVAD procedure. This led us to assume that there is considerable improvement in the dysregulation of plasma metabolism following the LVAD procedure. To determine whether LVAD can ameliorate the large polar metabolite imbalance observed in Patients with HF, we analyzed large polar metabolome changes using an untargeted metabolomics approach with a HILIC column. Following instrumental analysis, peak detection, and alignment, 9866 and 9561 features in positive and negative ion mode, respectively, were obtained in the metabolomic analysis.

First, the data quality of the features drawn from the metabolomic analysis was assessed as described in section 3.2. QC samples clustered together on the PCA score plots, suggesting that the metabolomic analyses were effective and reliable (Figure.S6). Then we did a preliminary verification of our hypothesis by PLS-DA analysis. The features of the metabolome from the three groups clustered in different regions of the PLS-DA score map (**Figure 4A, B**) revealed an overall difference in metabolite expression among them. Subsequently, we conducted pairwise comparisons between HCs and the preLVAD group, as well as between the preand postLVAD groups to analyze this metabolic change (as measured by OPLS-DA analysis, Figure S7). We identified 1628 metabolic characteristics that differed significantly between the HF and HC groups, and 837 metabolic characteristics that contributed significantly to distinguish the preLVAD and postLVAD groups, based on an FDR > 2 and *p* < 0.05 (Figure S8). Venn diagram analysis revealed 226 features that were significantly associated with LAVD-mediated improvement of HF (**Figure 4C**). Progenesis QI software screening annotated 153 of the metabolites and most were nonlipid polar metabolites (Table S3). To better understand the pathophysiological processes in LVAD-treated HF, we conducted a pathway enrichment analysis using these nonlipid polar metabolites. The results indicated that the cysteine and methionine, arginine and proline, and pyrimidine metabolism pathways were primarily implicated in the beneficial effects of LVAD (**Figure 4D**, Table S3). The heat maps for the pathway metabolic markers are shown in **Figure 4E**. In all, 15 markers from 12 pathways were visualized, including S-adenosylmethionine (SAM), L-methionine, dCMP, deoxycytidine, ADP, and 2-oxosuccinamate.

**Figure. 4.**
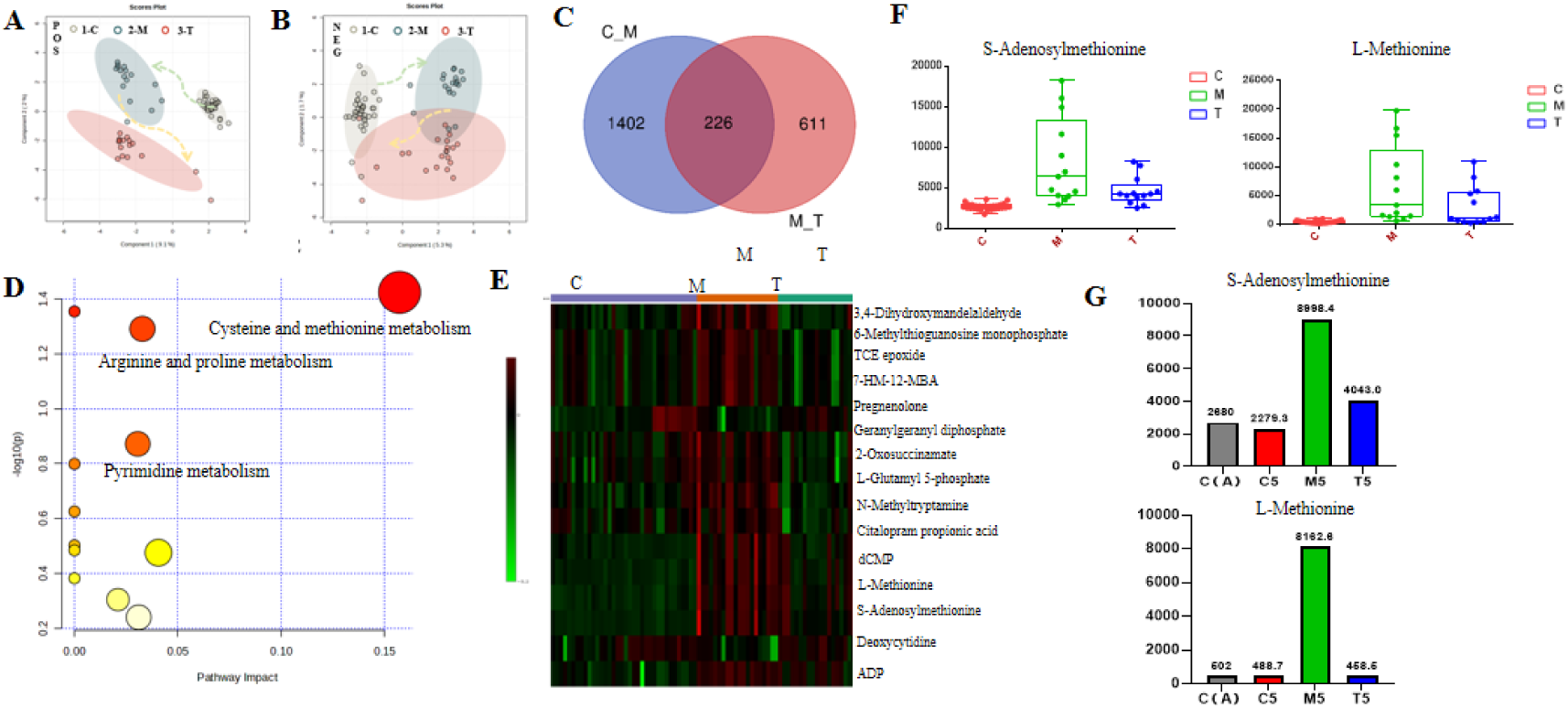
The effect of LVAD placement on the plasma amino acid metabolism (a-b: PLS-DA analysis in positive and negative mode; c: venn diagram of potential therapeutic substances for LVAD; d: pathway enrichment results of amino acid metabolites improved by LVAD; e: heat maps of LVAD- treatment amino acid metabolites from pathway enrichment results; f: box diagram of typical amino acids treated by LVAD; g:semi-quantitative analysis of S-adenosylmethionine and L-methionine in paired-individuals)

SAM and L-methionine levels showed prominent variations in the cysteine and methionine metabolic pathways. To determine the contribution of these two markers in ameliorating HF, we constructed box plots based on semiquantitative mass spectrometry data (**Figure 4F**). Both markers were up-regulated in the pre-LVAD plasma and were restored to healthy levels following LVAD pump implantation. To ensure these shifts were not artifactual, we made paired comparisons for each patient before and after the LVAD procedure (Figure.S9). The efficiency of SAM and L-methionine in the 20 Patients with HF was 70%. The typical change in the trends of these two markers in individual patients is shown in **Figure 4G**.

### LVAD increases the levels of PS (18:1/20:4) and canavaninosuccinate

Medical devices, such as LVADs, not only have therapeutic benefits, but are also associated with certain risks when implanted into the human body. To determine the influence of LVAD on peripheral plasma metabolism, we hypothesized that postLVAD causes some adverse metabolite disorders in Patients with HF. We compared the metabolome and lipidome data of healthy subjects (baseline) with that of postLVAD HF patients and identified 1349 metabolic characteristics that were significantly different. (**Figure 5A**, FDR>2, *p* < 0.05). Of these,10 lipids and 11 other metabolites were most markedly different (**Figure 5B**). A box diagram analysis revealed that PS (18:1/20:4) and canavaninosuccinate levels were significantly altered in the postLVAD group compared with the healthy control group (**Figure 5C**).

**Figure. 5.**
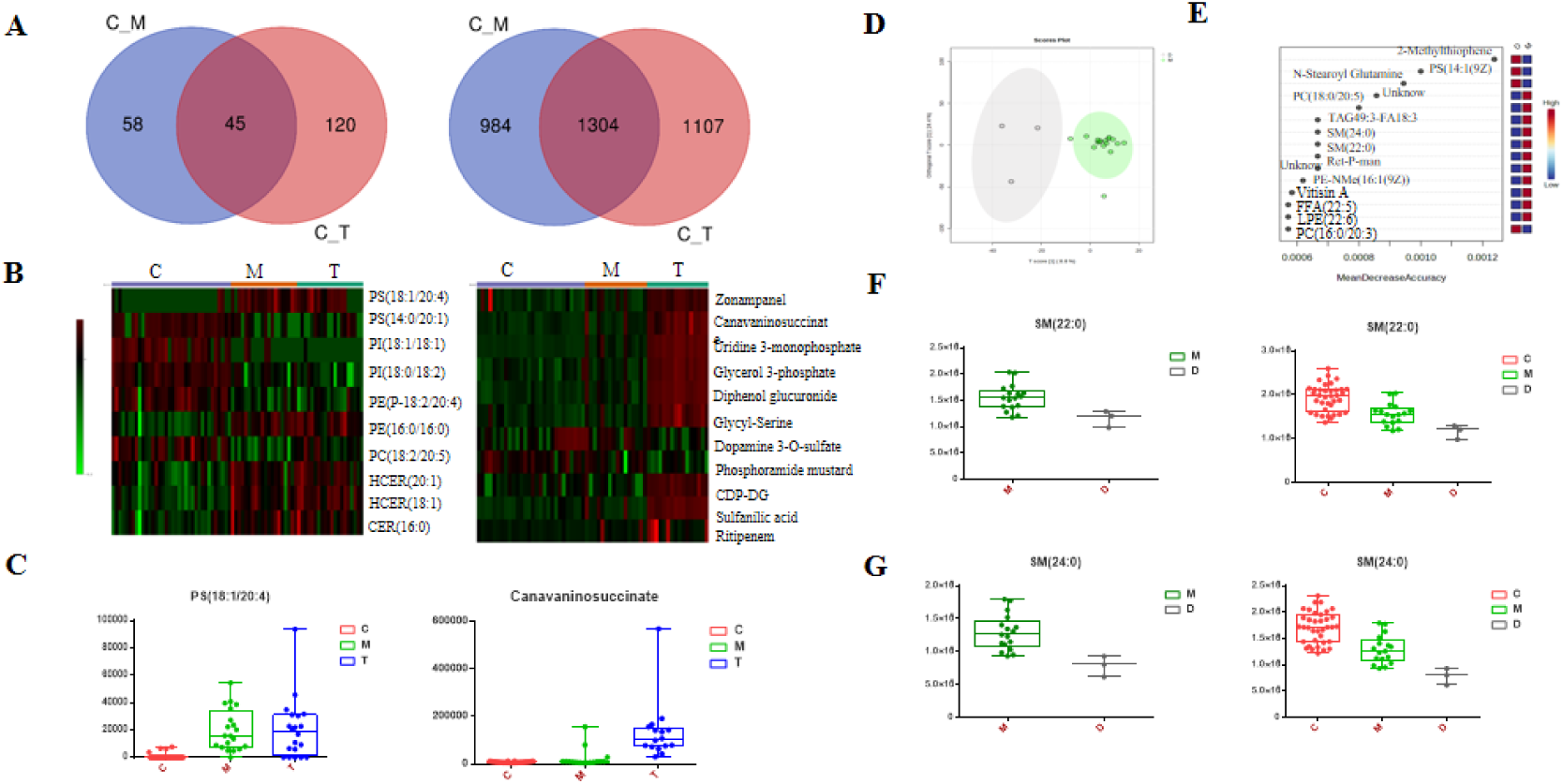
The dysregulated metabolites caused by LVAD placement and warning substances before loading pump (a: venn diagram of potential dysregulated substances after LVAD in lipidome and metabolome, respectively; b: heat maps of dysregulated lipids and metabolites caused by LVAD; c: box diagram of typical dysregulated substances caused by LVAD; d: OPLS-DA analysis of pre-LVAD plasma metabolome and lipidome between patients with a survival of less than one year and more than one year M: more than one year patients ; D: less than one year patients ;e: 15 key differential substances contributed to different survival years patients separated from random forest analysis; f-g: box diagram of typical abnormal metabolites in patients with a survival of less than one year)

### Low levels of preLVAD SM (24:0) and SM (22:0) are associated with death following LVAD installation

Three of the Patients with HF survived less than a year postLVAD implantation. We determined which metabolites were dysregulated in these patients, which could serve as markers for the effectiveness of LVAD postimplantation. The entire preLVAD plasma metabolome and lipidome data were compared between patients with a survival of less than one year with those surviving more than one year after LVAD placement (**Figure 5D**). Based on a random forest analysis, we identified 15 significantly different variables (**Figure 5E**). After assessing their biological significance and validation by semiquantitative analysis, two metabolites, SM (24:0) and SM (22:0), were confirmed to be abnormally present in patients who survived less than a year after LVAD placement compared with those surviving longer (**Figure 5F, G**). Although the significance of this result is limited because there were only three patients surviving less than a year, there was a notable decrease in the peripheral vein levels of these two SMs in all three patients.

### FFA (14:1), FFA (16:1), S-adenosylmethionine, and L-methionine are potential biomarkers for screening patients before LVAD installation

The previous analysis demonstrated that LVAD placement improves the levels of specific metabolites in HF patients (**Figure 2E, 4F**) and these changes are associated with the therapeutic effects of LVAD (**Figure 3, 4G**).To determine the clinical value of these findings, we calculated correlation coefficients for the individual metabolites [SAM, L-methionine, FFA (14:1), FFA (16:1), FFA (20:2), FFA (22:4), FFA (22:5), FFA (20:1), SM (24:0), SM (22:0)] and clinical indicators (BNP, LVEF). Although no metabolites were significantly associated with BNP, four [SAM, L-methionine, FFA (14:1), FFA (16:1)] exhibited a significant negative correlation with LVEF (**Figure 6A**). The differences in the levels of these four metabolites were confirmed by a receiver operating characteristic analysis to distinguish HF patients from HCs (AUC 0.7736–0.9931, all p < 2 × 10^−5^, **Figure 6B**).

**Figure. 6.**
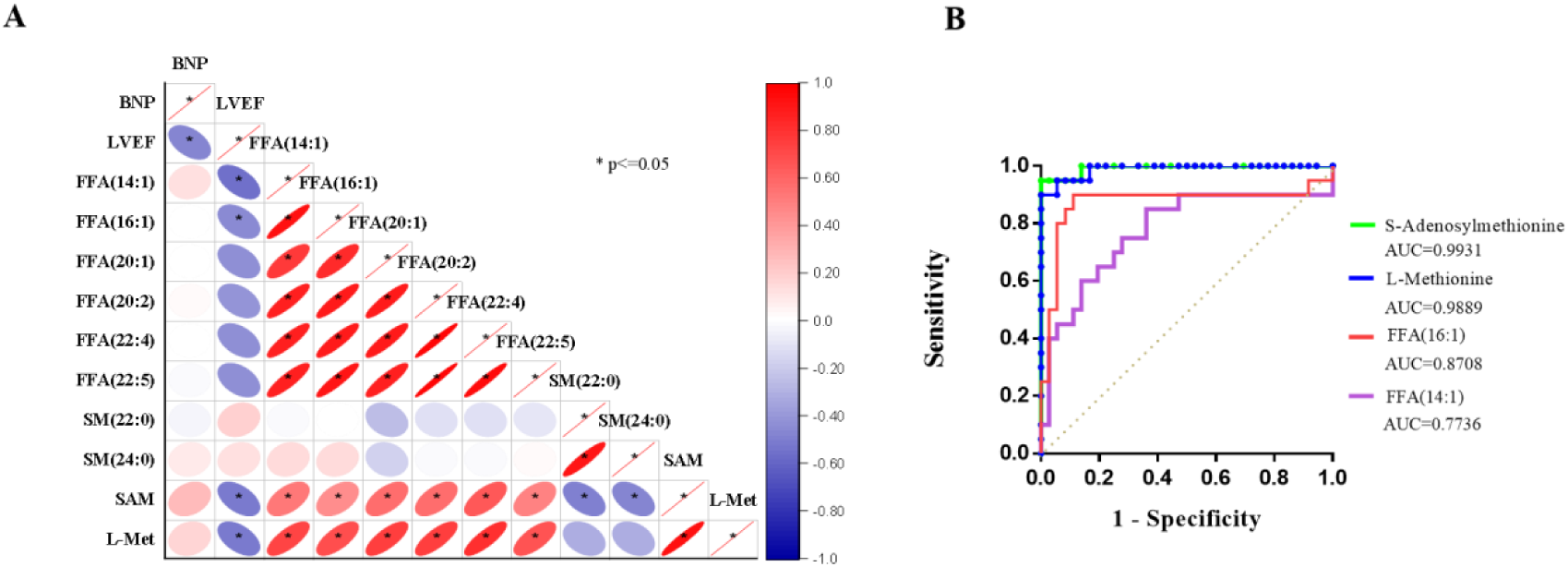
Correlation analysis and ROC curve analysis results

## Discussion

In this study, we successfully established an integrated pseudo-targeted lipidomic and untargeted metabolomic analysis method to identify changes in plasma metabolites in HF patients with LVAD. We found that 1) certain metabolites, amino acids, and lipids, including SAM, L-methionine, dCMP, deoxycytidine, ADP, 2-oxosuccinamate, FFA (14:1), FFA (16:1), FFA (20:2), FFA (22:4), FFA (22:5), and FFA (20:1) are dysregulated in preLVAD HF patients, but returned to normal levels postLVAD; 2) some metabolic pathways, including those of methionine, some lipids, and energy, were affected by LVAD treatment; 3) SAM, L-methionine, FFA (14:1), FFA (16:1), which exhibited significant negative correlations with LVEF, were primarily associated with the therapeutic effects of LVAD as determined by correlation and ROC analyses; and 4) postLVAD patients presented with some dysregulated metabolites and lipids compared with preLVADs, such as PS (18:1/20:4) and canavaninosuccinate. **Figure 7** shows a summary of our findings, which complement other LVAD studies ^[22, 23]^. Overall, our findings provide insight into the mechanism of LVAD-assisted HF improvement through the regulation of metabolism.

**Figure. 7.**
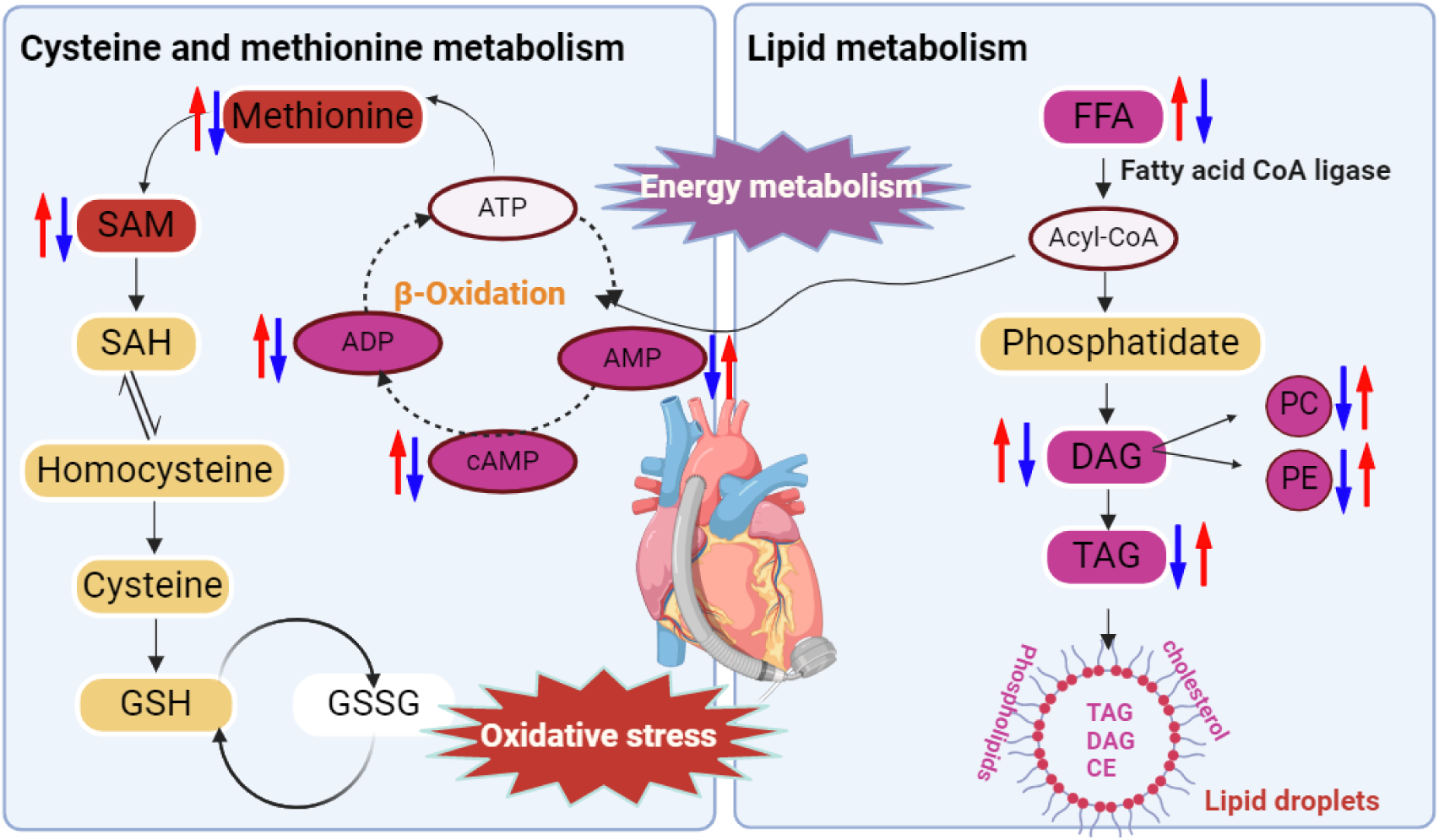
The potential mechanism of LVAD treatment

### Alterations of lipid metabolites

The most remarkable lipidomic changes observed in the postLVAD samples were FFAs. Six FFAs FFA (14:1), FFA (16:1), FFA (20:2), FFA (22:4), FFA (22:5), and FFA (20:1) were significantly increased in patients with HF, but were restored to normal levels after installation of the LVAD pumps with an average improvement rate of 81% in 20 patients. These results demonstrate that LVAD placement is associated with normalization in the abundance of FFAs in the peripheral circulation of patients with HF. FAs are considered the main source of heart fuel ^[24]^. FFAs, which are lipolysis by-products, provide 60%– 90% of the adenosine triphosphate required for myocardial metabolism under normal conditions ^[25]^. Higher plasma FFA concentrations were reported to be independently associated with a risk for HF. Increased FFA levels may result in increased triglyceride synthesis and fat storage, perhaps leading to lipotoxicity, cell apoptosis, and left ventricular dysfunction ^[26]^. Our results are consistent with previous studies and further show that LVAD contributes to the recovery of myocardial FFA uptake in patients with dilated cardiomyopathy.

### Alterations of amino acid metabolites

A variety of metabolites were attenuated following LVAD installation in patients with HF (**Figure 4E**), including SAM, L-methionine, dCMP, deoxycytidine, and ADP. Thus, LVAD helps ameliorate the disturbance in amino acid metabolism caused by HF. L-methionine is important for glutathione synthesis and lipid metabolism and counteracts oxidative stress ^[27]^. The inhibition of DNA damage and oxidative stress may represent effective treatment strategies for HF, since the chronic increase in oxygen-free radical production in the failing heart can result in a catastrophic cycle of mitochondrial DNA damage and functional decline ^[28]^. In addition, SAM is an important methyl donor to numerous transmethylation reactions, including DNA, RNA, catecholamines, and histones, as well as other proteins and phospholipids ^[29]^. dCMP and deoxycytidine are essential to DNA damage repair ^[30]^. Thus, we may infer that LVAD support reduces oxidative stress and increases DNA damage repair in the failing heart.

### Signaling pathways involved in postLVAD HF

Energy deficits, oxidative stress, and altered substrate utilization have been reported to contribute to the progression of HF ^[31]^. The heart is “metabolically flexible,” but mainly uses FAs to fuel mitochondrial oxidative phosphorylation and maintain ATP production ^[31, 32]^. In the present study, ADP and FFAs accumulated in HF plasma, which suggests that dysregulated energy metabolism contributes to HF (**Figure 2E, 4E**). After LVAD pump installation, the accumulated ADP and FFAs were consumed and their levels returned to a healthy state. These results suggest that LVAD support can restore energy to the failing heart. The measurement of energy metabolism-related substances preLVAD and postLVAD may assist in identifying the molecular mechanisms of LVAD-induced improvement in HF pathogenesis. Lipid peroxidation is a major contributor to oxidative stress ^[33]^. As expected, we observed increased oxidative stress indicators in the plasma of patients with HF, which is considered one of the major causes of congestive heart failure. Optimizing myocardial energy metabolism may serve as an additional approach for managing HF as our results indicate.

In the cysteine and methionine metabolism pathways, methionine and ATP are converted to SAM by methionine adenosyl transferase (MAT) ^[34]^. The methionine– homocysteine cycle has been implicated in methylation and redox balance via regulating the synthesis of the antioxidants, cysteine, and glutathione, and controlling the amount of S-adenosyl homocysteine and SAM ^[35]^. Homocysteine (Hcy), a metabolite of methionine, is a risk factor for HF and is correlated with left ventricular mass and wall thickness ^[36, 37]^. In the present study, a marked change in SAM and L-methionine levels were observed between the pre and postLVAD groups. Furthermore, dCMP and ADP levels in the cysteine and methionine metabolism pathway were significantly altered following LVAD treatment, which suggests that LVAD contributes to the changes in oxidant status and affects methylation reactions by altering the flux through the methionine–homocysteine cycle.

### Identification of novel biomarkers and their diagnostic value

Correlation and ROC analyses were done to identify candidate diagnostic markers and evaluate their clinical value. No metabolites were significantly associated with BNP, whereas four SAM, L-methionine, FFA (14:1), and FFA (16:1) were negatively correlated with LVEF (**Figure 6A**). Plasma BNP is a guideline-mandated diagnostic biomarker widely used for HF; however, the efficacy of using BNP measurements for HF treatment remains unclear ^[38]^. LVEF is also a clinical parameter used to describe HF with LVEF in 40%–50% of cases ^[39]^. In the present study, more metabolites were correlated with LVEF rather than BNP, which suggests that the structural dysfunction of the left ventricle may have an important role in the perturbance of the plasma metabolome in patients with HF. The metabolites in our study are likely to provide strong clues for studying the typing and diagnosis of HF. Univariate ROC analysis confirmed the diagnostic efficacy of these markers, with three metabolites exhibiting AUC values greater than 0.8 (**Figure 6B**), (AUC values: SAM: 0.9931, L-methionine 0.9889). In addition, the levels of these biomarkers significantly improved after loading the pump, indicating that their detection in the peripheral vein, which is readily accessible, may help screen patients requiring LVAD therapy. On the other hand, SM (24:0) and SM (22:0), were abnormal in patients who survived less than one-year postLVAD, indicating that may be useful biomarkers to guide treatment. Future studies should focus on developing readily available, noninvasive clinical measures for HF in patients before and after LVAD.

### Identification of potentially adverse substances

Changes in PS (18:1/20:4) and canavaninosuccinate were significantly perturbed in the postLVAD group (**Figure 5C**). Experimental studies have suggested that PS (18:1/20:4) disorder is associated with Alzheimer’s disease ^[40]^, aging, and traumatic brain injuries ^[41]^. Canavaninosuccinate is elevated in hepatopathy progression ^[42]^. These findings suggest that LVAD may have side effects in the brain or liver, thus translational studies should focus on these aspects, and determine their effects on neuroinflammation or liver injury. This may lead to the development of targeted adjuvant drugs to improve survival following pump installation.

### Limitations

Our study has several limitations. First, the study is exploratory and preliminary based on the discovery cohort. Only 20 patients with HF were recruited because of the need for specificity of the treatment protocol as well as time constraints. Second, the levels of the various metabolites and lipids provided were determined by relative quantitation. Validation studies with absolute quantitation of the proposed markers in a larger number of postLVAD HF patients are needed before these candidate markers can be applied to actual clinical practice. Third, we concluded that molecules whose levels were found altered were specific to LVAD. Studying the association of these biomarkers with many other diseases and treatments was difficult because of various practical problems, such as the nonavailability of patient samples at the appropriate times. Further evaluation of the biomarkers with respect to other disease conditions is required.

## Conclusion

In summary, we used the innovative methodology of integrated pseudo-targeted lipidomic and untargeted metabolomic analyses to comprehensively identify metabolic changes in ischemic patients with HF undergoing LVAD treatment. We examined the physiological mechanism of LVAD for treating HF through the regulation of FAs and methionine levels. Based on our findings, we propose that oxidative stress and energy metabolism dysregulation may occur in patients who develop HF postLVAD. SAM, L-methionine, FFA (14:1), and FFA (16:1), are potential diagnostic markers for HF or may predict LVAD efficacy. In three patients who died within a year postLVAD, we observed decreases in SM (24:0) and SM (22:0) immediately before LVAD installation, which suggests that a decrease may serve as a warning sign against LVAD. Furthermore, we demonstrated that PS (18:1/20:4) and canavaninosuccinate were significantly perturbed in postLVAD patients. Finally, this work provides an impetus to further study the molecular mechanisms related to LVAD and to evaluate potential biomarker panels for HF diagnosis or LVAD prognosis in clinical practice.

## Data Availability

Data availability statement: All relevant data are within the manuscript and its Additional files.

## Non-standard Abbreviations and Acronyms

AUC: Area under the curve
BNP: B-type natriuretic peptide
FA: Fatty acids
HC: Healthy controls
HF: Heart failure
IS: Internal standards
LVAD: Left ventricular-assist device
LVEF: Left ventricular ejection fraction
PCA: Principal component analysis
QC: Quality control
ROC: Receiver operating characteristic curve

## Acknowledgments

The authors thank Dr. Jian Xu and Dr. Yi Ding for valuable suggestions. The authors thank Dr. Lingyu Han for help with blood sample collection and Ms. Weihua Wang and Mr Yu Tian for help with lipidomics analysis.

## Author contributions

Na Zhang and Hao Chen designed the experiments and edited the manuscript. Yu Tian developed the lipidomics and metabonomics method and performed the data analysis. Heping Li participated in the discussion of the experimental plan and confirmed the parameters.XiaoYu Xu designed and completed the graphical abstract. Xuman Zhang and Zhifu Han were responsible for the LVAD product. Haitao He and Guowei He were responsible for collecting blood samples from the patients before and after LVAD implantation. Yu Zhang was responsible for collecting blood samples from health volunteers and also led the entire project.

